# Understanding the barriers to pooled SARS-CoV-2 testing in the United States

**DOI:** 10.1101/2021.04.17.21255440

**Authors:** Eli P. Fenichel, Anna Gilbert, Gregg Gonsalves, Anne L. Wyllie

## Abstract

Pooled testing for SARS-CoV-2 detection is instrumental for increasing test capacity while decreasing test cost, key factors for sustainable, long-term surveillance measures. While numerous pooled approaches have been described, uptake by labs has been limited. We surveyed 90 US labs to understand the barriers to implementing pooled testing.

---

Early in the pandemic testing for SARS-CoV-2 emerged as an Achilles’ heel of the United States’ national response (1). The failure to scale-up testing programs rapidly, delays in test processing and return of results led to delays downstream in self-isolation, diagnosis and treatment, undercounting of infections, fear and confusion. Proposals for novel strategies involved pooling of samples from multiple individuals into one testing run. Many countries, including Israel, Germany, South Korea and China, rapidly implemented pooling as part of national plans. The US never implemented pooling as part of its own strategy in earnest (2). According to Google Scholar, over 10,000 papers with key words “COVID” and “pooling” were ‘published’ in 2020. Pooling samples to increase throughput is not a new idea (3). For most realistic test positive levels even simple pooling designs greatly increase capacity (4). So, why has pooled testing been so rare in the US? To resolve this question, we explored the barriers facing labs to expand testing capacity.

On December 9, 2020, we invited 362 labs that had contacted the Yale School of Public Health expressing interest in implementing the SalviaDirect test (5), to participate in a survey to evaluate testing constraints and pooling strategies for SARS-CoV-2 testing. The survey was distributed using Qualtrics and three reminders were sent. The survey closed on January 21, 2021. Of the 93 responses received (25.7% response rate), 90 responses were from CLIA certified labs conducting SARS-CoV-2 testing. The remaining three were excluded from the analyses. Nearly half of the labs responding were for profit (n=42), followed by university affiliated labs (n=9), with the remainder made up by community, non-profit, and government laboratories. The respondent labs reported serving a variety of testing populations, the bulk consisting of outpatient and community testing. While most conducted diagnostic testing, many labs reported testing also for general surveillance and screening for specific events. The high reporting of diagnostic testing, which is likely representative across all CLIA labs, is important in the context of pooling, which differs from testing for screening and surveillance at the population scale.

The survey included an open-ended question about the barriers to pooling. A major common barrier to the implementation of pooling by labs was a lack of methods accepted by the necessary authorities, including the FDA, CLIA regulations or their own laboratory directors. Reports of a lack of clear protocols and guidance on the methodology for pooled testing contributed to this. Many labs expressed that even if protocols were available, a lack of time and resources limited their ability to validate these in-house, preventing proof-of-concept implementation. Thus, demonstration of the effectiveness of pooling in their setting is lacking. In that regard, numerous labs reported that local case-positivity rates were simply too high to warrant pooling, despite recent lab-based data demonstrating the benefit of pooling five samples up to a ∼30% test positive rate (4). Throughout, there were a substantial number of concerns about the effect of pooling on the sensitivity of detection and the increased risk for false negative results due to sample dilution.

Operational and administrative barriers were also a common theme. Labs frequently reported that reflex testing of the most common Dorfman approach (3), where samples from positive pools are re-visited for individual re-testing, are too logistically difficult to manage or too resource demanding. Labs viewed the need to re-visit samples of positive pools as disruptive to the standard testing flow and staffing practices, expressing concerns that this would add to the time of reporting out positive tests. A major concern was the additional logistics required for following samples through the pooled testing process and the lack of the necessary software to track samples through the workflow. Important to any testing program were concerns regarding reimbursement and billing when samples are tested in pools as well as contract obligations. Limited staff to dedicate to tackling these issues came up repeatedly.

Survey responses demonstrated that the SARS-CoV-2 testing environment in the US is highly heterogeneous, particularly in regards to the supplies, testing platforms and the variety of sample types received. While a majority of labs use 96-well PCR instruments, 384-well instruments are also common, with some labs reporting non-standard plate sizes in upstream processes. Many labs utilize multiple instruments in their testing procedures, which can vary in the number of samples each can process. These inter-lab variations in testing workflows are important for pooled testing strategies: a protocol developed for one lab may not be easily translated into another. Those developing pooled testing strategies need to recognize that labs are working with fixed physical and human capital constraints, and that new labs are not being built around pooling strategies, such constraints cannot be assumed away.

Higher test throughput, lower costs per test, and faster turnaround times are all margins for testing improvement. Increasing the number of samples per test in a pooling structure can be part of that solution. Laboratories using an extraction process report that it takes on average 4.6 hours to test a sample from start to finish (extraction-less = 3.0 hours), but there is substantial variation (Figure 1). The small number of labs (n=6) in our sample that engage in pooling reported total testing time inline with these figures. On average samples wait in labs for 93 minutes prior to extraction-less workflows and 279 minutes prior to extraction-based tests. The average time to set-up and run PCR is 94 minutes, with little difference between labs. For labs performing full RNA extraction, this process takes on average 57 minutes. Reset or cycle times between processes are on average 18 minutes for extraction and PCR. These times are one motivation for laboratories focused on faster turn around to be wary of designs that require retesting of samples, which can add hours to the process.

**Figure 1.**
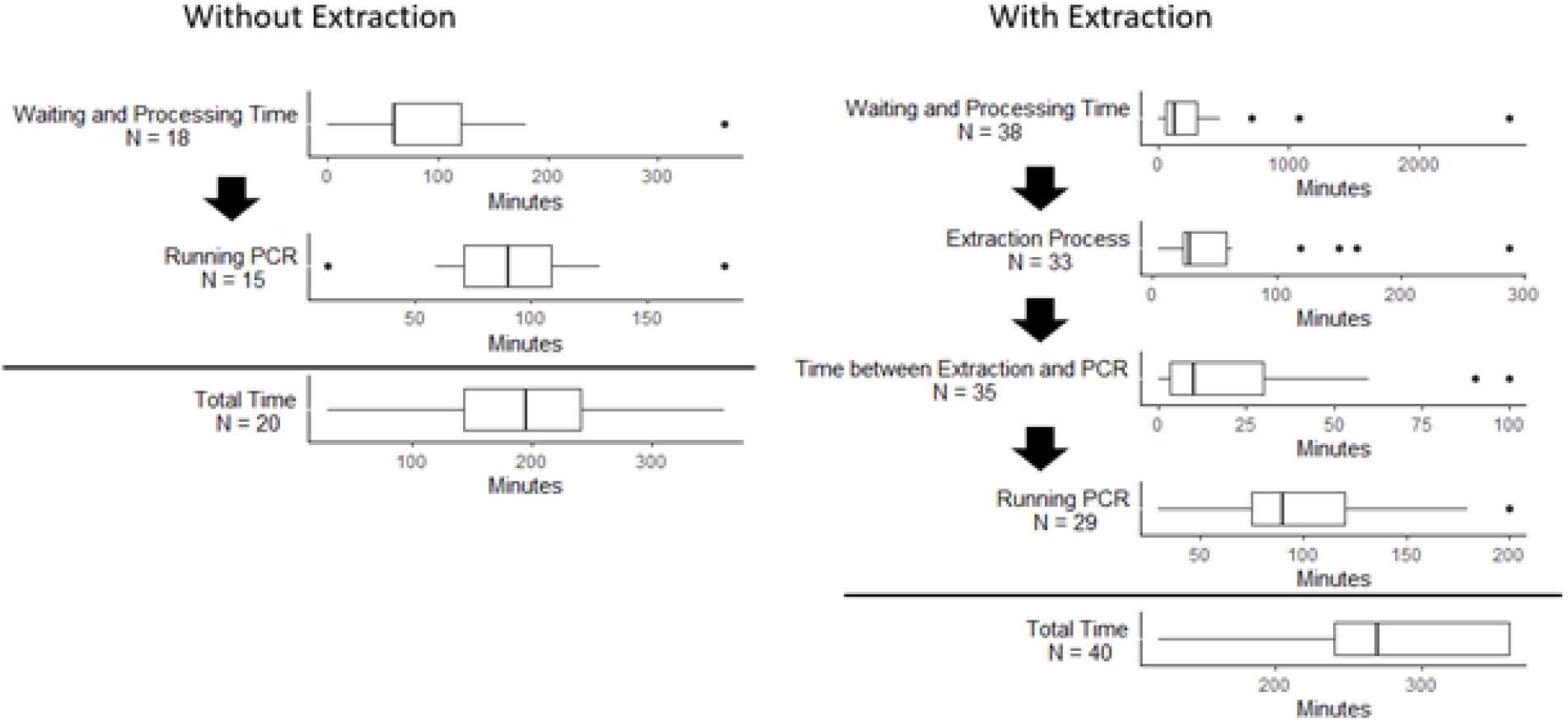
The variation in timing constraints for critical steps of SARS-CoV-2 testing workflows with or without RNA extraction.

The need for testing will remain for the years to come. Pooled testing offers sustainable surveillance measures that support long term programs, essential for the early detection of virus resurgence or the emergence of variants of concern. While testing programs can be maintained by pooled PCR testing, labs require guidance on how to transition from traditional diagnostic testing to pooled surveillance. Importantly, our survey suggests that the major barriers to uptake and implementation of pooled testing in the US may not simply be the number of tests a lab can process per day, rather it is the lack of adequate resources guidance on clinical best practices to transition to pooling. Additionally, a number of labs did not feel that pooled testing was necessary given current testing capacity constraints at present. If the appropriate resources were made available however, labs reported that even with current non-pooled testing strategies, they could on average already increase testing capacity by 60%. While labs need to see more evidence supporting the ability of pooled testing strategies to successfully move from theory to a reliable and resource-savings laboratory practice, logistical solutions to support implementation and general processing remain vital. Responses to our survey highlight the importance of consulting end-users, those that solutions are being designed for, so challenges can be crafted to meet the specific needs out in the field. It may be surprising to those designing pooling strategies to learn that labs view pooling as more time consuming, delaying test reporting.

## Data Availability

Data available on request from the authors

## References

1. Schneider EC. Failing the Test - The Tragic Data Gap Undermining the U.S. Pandemic Response. N Engl J Med. 2020 Jul 23;383(4):299–302.

2. Wu KJ. Why Pooled Testing for the Coronavirus Isn’t Working in America. The New York Times [Internet]. 2020 Aug 18 [cited 2021 Feb 21]; Available from: https://www.nytimes.com/2020/08/18/health/coronavirus-pool-testing.html

3. Dorfman R. The Detection of Defective Members of Large Populations. Ann Math Stat. 1943;14(4):436–40.

4. Watkins A, Fenichel E, Weinberger D, Vogels CBF, Brackney D, Casanovas-Massana A, et al. Increasing SARS-CoV-2 Testing Capacity with Pooled Saliva Samples. Emerging Infectious Disease journal [Internet]. 2021;27(4). Available from: https://wwwnc.cdc.gov/eid/article/27/4/20-4200_article

5. Vogels CBF, Watkins AE, Harden CA, Brackney DE, Shafer J, Wang J, et al. SalivaDirect: A Simplified and Flexible Platform to Enhance SARS-CoV-2 Testing Capacity. Med [Internet]. 2020 Dec 26; Available from: http://www.sciencedirect.com/science/article/pii/S2666634020300763

